# Excess GDNF associates with a subtype of schizophrenia with enhanced dopamine and increased prefrontal dysfunction

**DOI:** 10.1101/2025.08.11.25333355

**Authors:** Isabel Runneberger, Daniel R. Garton, Peyman Choopanian, Ana R Montaño-Rodríguez, Mehdi Mirzaie, Funda Orhan, Vladislav Yakimov, Joanna Moussiopoulou, Vilma Iivanainen, Soophie Olfat, CDP-Working group, Andrea Schmitt, Peter Falkai, Salla E Keskitalo, Markku T Varjosalo, Simon Cervenka, Carl M Sellgren, Sophie Erhardt, Fredrik Fagerström-Billai, Anastasios Damdimopoulos, Göran Engberg, Elias Wagner, Florian J. Raabe, Jaan-Olle Andressoo

**Affiliations:** Division of Neurogeriatrics, Department of Neurobiology, Care Sciences and Society (NVS), Karolinska Institutet; Stockholm, Sweden; Department of Pharmacology, Faculty of Medicine, Neuroscience Center & Helsinki Institute of Life Science, University of Helsinki; Helsinki, Finland; Department of Pharmacology, Faculty of Medicine, University of Helsinki, Helsinki, Finland; Department of Physiology and Pharmacology, Karolinska Institutet, Stockholm, Sweden; Department of Psychiatry and Psychotherapy, LMU Hospital, LMU Munich, 80336 Munich, Germany; International Max Planck Research School for Translational Psychiatry (IMPRS-TP), 80804 Munich, Germany; Max Planck Institute of Psychiatry, 80804 Munich, Germany; Evidence-based Psychiatry and Psychotherapy, Faculty of Medicine, University of Augsburg, Augsburg, Germany (CDP working group); Department of Psychiatry, Psychotherapy and Psychosomatics, Medical Faculty, University of Augsburg, BKH Augsburg, Augsburg, Germany. (CDP working group); German Center for Mental Health (DZPG), partner site Munich-Augsburg; Laboratory of Neurosciences (LIM-27), Institute of Psychiatry, University of São Paulo (USP), São Paulo-SP 05403-903, Brazil; Institute of Biotechnology, Helsinki Institute of Life Science HiLIFE, University of Helsinki, Helsinki, Finland; Helsinki Proteomics Center, University of Helsinki, Helsinki, Finland; Department of Biochemistry and Developmental Biology and Translational Cancer Medicine Program, Faculty of Medicine, University of Helsinki, Helsinki, Finland; Department of Medical Sciences, Psychosis Research and Preventive Psychiatry, Uppsala University, Uppsala, Sweden; Centre for Psychiatry Research, Department of Clinical Neuroscience, Karolinska Institutet & Stockholm Health Care Services, Region Stockholm, Sweden; Bioinformatics and Expression Core Facility, Karolinska Institutet, Stockholm, Sweden; Department of Medicine, Huddinge, Karolinska Institutet, Stockholm, Sweden; Institute of Sport Science and Innovations, Lithuanian Sports University, Kaunas, Lithuania

## Abstract

Schizophrenia (SCZ) displays a heterogeneous etiology. In about half of the patients, first-episode psychosis (FEP) associates with increased levels of striatal dopamine (DA), while cognitive and negative symptoms associate with reduced DA function in the prefrontal cortex (PFC). However, the mechanism underlying these changes in DA is unclear. Abnormal increase in glial cell-line derived neurotrophic factor (GDNF) leads to similar SCZ-like symptoms in mice, but its relevance in patients has remained uncertain. Here, we use omics to examine three patient cohorts and identify a subgroup of patients with elevated GDNF. High GDNF associates with increased DA in cerebrospinal fluid, reduced DA-related PFC gene expression, aggravated negative SCZ symptoms, and a specific serum proteomics pattern. Post-mortem analysis revealed a GDNF-high specific striatal gene expression with significant overlap between mice and patients. Collectively, our data suggest a mechanism for DA imbalance in a subgroup of patients with SCZ.

## INTRODUCTION

Schizophrenia (SCZ) is a severe neuropsychiatric disorder with a prevalence of about 1%, and with high individual as well as societal burden^1,2^. Despite decades of research revealing highly polygenic architecture and clinical heterogeneity of SCZ, including about 30-50% discordance between monozygotic twins^3–7^, its molecular mechanisms remain poorly understood. Current evidence suggests that an increase in striatal dopamine (DA) drives positive symptoms such as hallucinations and delusions, while a reduction in DA in prefrontal cortex (PFC) contributes to cognitive impairment and negative symptoms such as apathy and avolition^3,8–10^. Supporting this, most antipsychotic medication acts via pharmacological blocking of DA receptors, with the main focus on DA receptor 2 (D2R)^8,11,12^, and conversely, drugs enhancing DA function increase the risk for psychosis^13–15^. However, only about 50 to 70% of patients respond to treatment with DA receptor antagonists, and cognitive and negative symptoms lack specific treatments altogether^8,16–19^. Illustrating the role of excess striatal DA in psychosis, brain imaging studies have revealed that increased DA synthesis and release in the striatum (STR) is observed in about 50% of individuals with primary first-episode psychosis (FEP), and patients with FEP with elevated striatal DA are almost exclusively the patients who respond to DA-blocking anti-psychotics^9,11,12,20^. However, the mechanism underlying increased striatal DA and reduced PFC DA in SCZ remains elusive and is thereby limiting causal treatment design. Illustrating a need for finding new knowledge-based and potentially more selective treatment targets tailored for patient sub-groups, pharmacological intervention by blocking DA receptors elicits a wide range of side effects such as those associated with tuberoinfundibular dopamine circuitry, which innervates median eminence controlling prolactin release from the pituitary^8^.

We recently discovered that the levels of glial cell line-derived neurotrophic factor (GDNF) – a secreted protein predominantly expressed by the striatal interneurons^21–23^ and a potent enhancer of nigrostriatal DA function^22,24–28^ – are elevated in the cerebrospinal fluid (CSF) of primary FEP patient cohort from Karolinska Schizophrenia Project (KaSP, Sweden) and in post-mortem striata from patients with SCZ obtained at the University of Pittsburgh (USA)^29,30^. In both cases, about 20% of patients displayed GDNF levels up to about two-fold higher than seen in any of the control individuals^29^. In mice, we found that a similar about two-fold increase in endogenous GDNF expression results in a range of SCZ-like brain circuit alterations, including enhanced striatal DA and reduced DA in the PFC, with related features such as apathy, polydipsia, and defect in pre-pulse inhibition test, features which associate with SCZ, demonstrating that in mice, endogenous GDNF increase is sufficient to induce SCZ-like condition^29^. However, whether excess GDNF observed in about 20% of patients^29^ triggers or contributes to SCZ development in that patient subset remained unknown. To address this, we analyzed global gene expression in individual patients’ post-mortem STR and PFC, evaluated patients with FEP for the Positive and Negative Syndrome Scale (PANSS) and the Clinical Global Impression (CGI) scale, measured levels of neurotransmitters and other neuronal function regulators in the CSF of patients with FEP, analyzed serum proteomics in patients with FEP, and performed comparative analysis with animal model with increased expression of endogenous GDNF.

## RESULTS

### Elevated striatal GDNF expression in post-mortem brains of patients with SCZ

To explore striatal transcriptomic alterations in SCZ, we analyzed the raw microarray data from the post-mortem STR of patients with SCZ and unaffected healthy controls (HCs)^30^. We found 434 differentially expressed genes (DEGs; FDR < 0.1) in individuals with SCZ compared with HCs; 160 upregulated genes and 274 downregulated genes (Fig. 1a, list of genes in Extended Data Table 1). We found that *GDNF* is among the upregulated genes (Log_2_FC = 0.252, FDR = 0.0737). For the analysis of relative expression levels of GDNF, we transformed the microarray data (Extended Data Fig. 1a) into RNAseq-comparable data^31^ and normalized the median of HCs to one (Fig. 1b). Thereby, we could reveal that seven out of eighteen patients with SCZ showed a particularly high expression of GDNF (SCZ GDNF-high) compared with healthy controls (P = 8.38e^−7^), whereas the remaining eleven patients (SCZ GDNF-normal) did not have increased GDNF expression compared with HCs. Since GDNF is a strong enhancer of nigrostriatal DA function^22,24–28^, this result indicates a potential subgroup-specific biology and prompted us to perform further analysis.

**Fig. 1.**
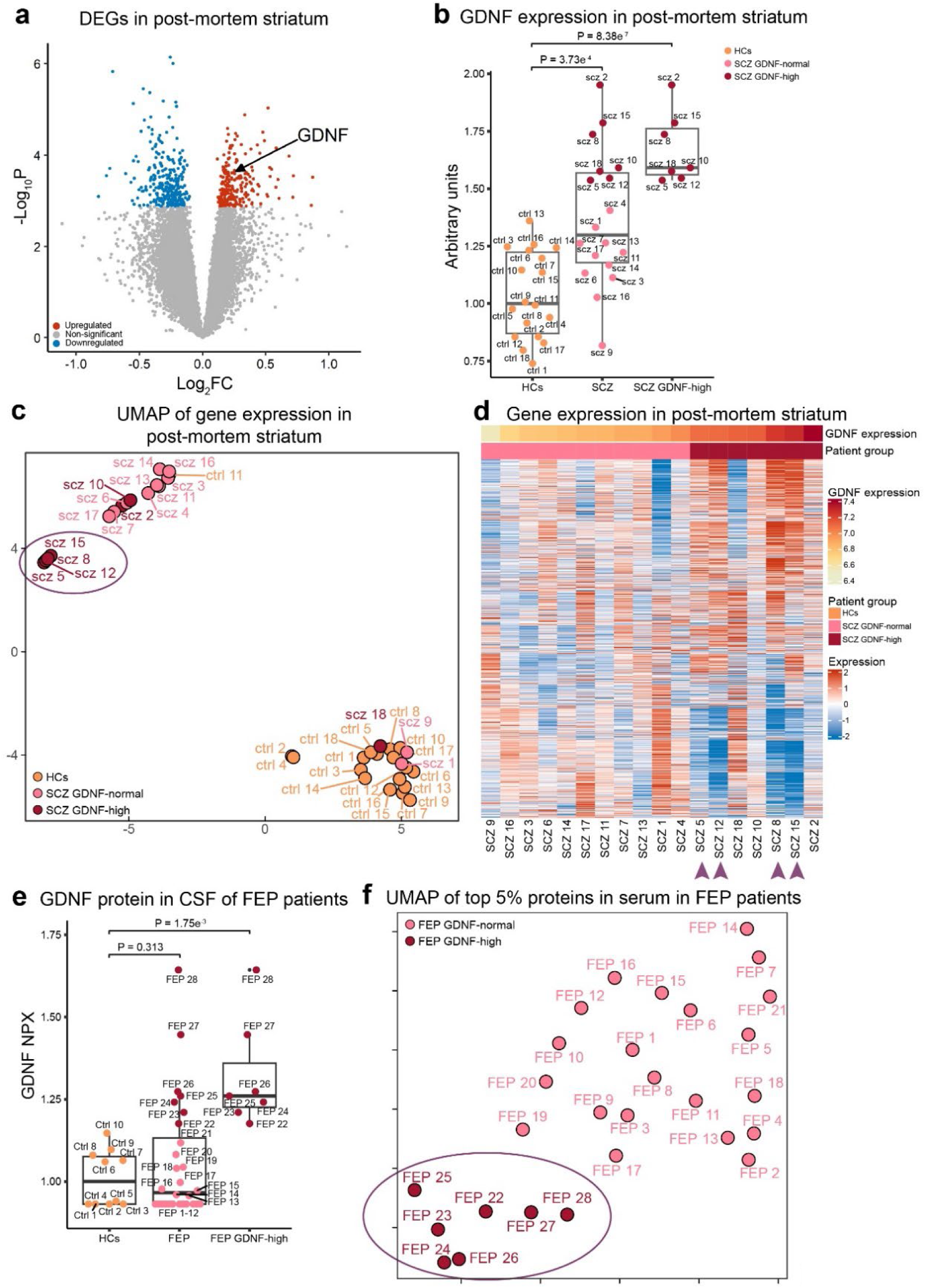
GDNF expression in CSF and post-mortem STR of patients with SCZ and a patient subgroup gene expression pattern. (**a**) Gene expression in STR in patients with SCZ compared with HCs, based on raw microarray data of postmortem patient samples^30^. Significantly regulated genes (P < 0.1) in red (upregulated) and blue (downregulated). GDNF is upregulated. (**b**) GDNF expression levels in post-mortem STR in patients with SCZ (N=18) compared with HCs (N=18), with GDNF-high patients (N=7) also shown separately. GDNF is significantly upregulated in both the whole SCZ patient group (P = 3.73e^−4^; Welch’s t-test) and the GDNF-high patient group (P = 8.38e^−7^; Welch’s t-test) compared with HCs. Microarray data has been transformed into RNAseq-comparable data using formula 1.31*m-7.475^31^ and normalized to median_ctrl_ = 1. (**c**) UMAP based on gene expression in post-mortem STR of patients with SCZ and HCs, supervised by disorder. A subgroup of four GDNF-high patients (SCZ 5, 8, 12 and 15) clustered separately (circled). (**d**) Striatal gene expression pattern in post-mortem SCZ patients. The four GDNF-high patients from (c) exhibit a different gene expression pattern than the rest of the patients with SCZ. (**e**) GDNF protein levels in CSF in a live patient cohort of patients with FEP from LMU Hospital, compared with HCs and patients with high GDNF are also shown separately. GDNF shows a trend towards upregulation in the whole patient cohort (P = 0.313; Welch’s t-test) and is significantly upregulated in the GDNF-high patients (P = 1.75e^−3^; Welch’s t-test). (**f**) UMAP based on protein analysis in serum in patients with FEP, using the top 5% proteins from AUC and components describing 90% variance from PCA. Patients with FEP with high GDNF in CSF cluster separately and are circled.

### Stratification of SCZ subtypes based on striatal gene expression

Uniform Manifold Approximation and Projection (UMAP) analysis of the post-mortem patients’ striatal gene expressions revealed three different clusters (Fig. 1c). One major cluster (Fig. 1d, bottom right corner) mainly consisting of HCs, another major cluster (upper left corner) with primarily patients with SCZ, and a small separate cluster of four patients with SCZ (SCZ 5, 8, 12, and 15) – all with high GDNF. Investigating the gene expression patterns, these four GDNF-high patients with SCZ displayed different gene expression patterns compared with the rest of the patients (Fig. 1d). Moreover, differential gene expression analysis of these four SCZ patients compared with healthy controls revealed 4791 upregulated genes and 5127 downregulated genes (FDR < 0.05; list of genes in Extended Data Table 2), illustrating the separate biology of a distinct subtype of SCZ characterized by high *GDNF* gene expression.

### GDNF in CSF and serum proteomics analysis in an independent FEP patient cohort

Previously, we reported up to about two-fold increase in GDNF in cerebrospinal fluid (CSF) in the Swedish Karolinska Schizophrenia Project (KaSP) FEP patient cohort, with a clear increase of GDNF above the highest levels in HCs observed in about 20% of patients with FEP^29^. To further explore our previous finding, we next investigated GDNF protein levels with proximity extension assays (PEA) by OLINK Bioscience in CSF in an independent FEP patient cohort collected in Germany at the Ludvig-Maximillians University (LMU) Hospital and found that seven out of twenty-eight patients (25%) had higher levels of GDNF than any of the HCs (Fig. 1e; P = 1.75e^−3^), similar to what was reported in the KaSP cohort of patients with FEP^29^. The average GDNF level across all patients with FEP did not differ compared to HCs (Fig. 1e; P = 0.313). Proteome analysis of serum from the LMU FEP patient cohort using LC-MS/MS identified a total of 485 proteins. For each protein, we performed a univariate analysis and calculated the area under the curve (AUC) for the receiver operating characteristic (ROC) curve to separate patients from HCs. Conducting a principal component analysis (PCA) with the top 5% proteins with the highest AUC (24 proteins; Extended Data Fig. 1b), we found separate clustering of patients with FEP with high GDNF levels in CSF (Extended Data Fig. 1c). Selection of the top components explaining 90% of the PCA and application of these in a UMAP further improved separation of GDNF-high FEP patients from GDNF-normal FEP patients (Fig. 1f). Additionally, we implemented a multi-class ROC analysis using a support vector machine (SVM) model using these 24 proteins and observed perfect classification accuracy with AUC=1 (Extended Data Fig. 1d), indicating that protein biomarkers in serum from patients with FEP could be used to identify patients with high CSF GDNF expression.

### Analysis of striatal Gdnf expression in GDNF conditional Hypermorph mouse model

Previously, we developed a conditional Hypermorph mouse model of GDNF (*Gdnf^cHyper^*), which allows for conditional upregulation of endogenous GDNF expression via conditional 3’UTR replacement^29^. The animals have one or two copies of the *Gdnf*^cHyper^ allele (Fig. 2a), which contains a FLEx cassette with bovine growth hormone polyadenylation sequence (bGHpA) in an inverted position, placed between the stop codon and the natural 3’UTR of the *Gdnf*. Upon Cre-mediated recombination, bGHpA; a 3’UTR sequence which elevates mRNA and protein levels^32^, is reverted to the correct position and will terminate transcription before the natural 3’UTR, resulting in increased GDNF expression in cells which naturally transcribe *Gdnf* (Fig. 2a, b). Here, we use RNAseq data from STR from the *Gdnf^cHyper^*;Nestin-Cre mouse model^29^ (available at GEO: GSE162974) to investigate GDNF-dependent gene expression profiles. Differential gene expression analysis yielded 1311 DEGs for *Gdnf^wt/cHyper^*;Nestin-Cre, and 288 for *Gdnf^cHyper/cHyper^*;Nestin-Cre mice, (FDR < 0.1; list of genes in Supplementary table 3). Upregulation of Gdnf in the *Gdnf^cHyper^*mice has previously been confirmed with rt-qPCR^29^, but analysis of RNAseq data allows for investigation of individual exons and the 3’UTR of the gene. An allele dose-dependent increase was observed in expression of the mature protein-coding exon 3 of Gdnf in *Gdnf^cHyper^*;Nestin-Cre animals (Fig. 2c, Extended Data Fig. 2a). Heterozygous *Gdnf^wt/cHyper^*;Nestin-Cre mice show about two to three times increase in mean Gdnf exon 3 expression compared with wild type (WT), whereas homozygous *Gdnf^cHyper/cHyper^*;Nestin-Cre animals have mean expression levels of four to six times higher than WT. Conversely, we observe that the expression of the natural Gdnf 3’UTR is allele dose-dependently downregulated in the *Gdnf^cHyper^*;Nestin-Cre animals as expected (Fig. 2c, Extended Data Fig. 2a).

**Fig. 2.**
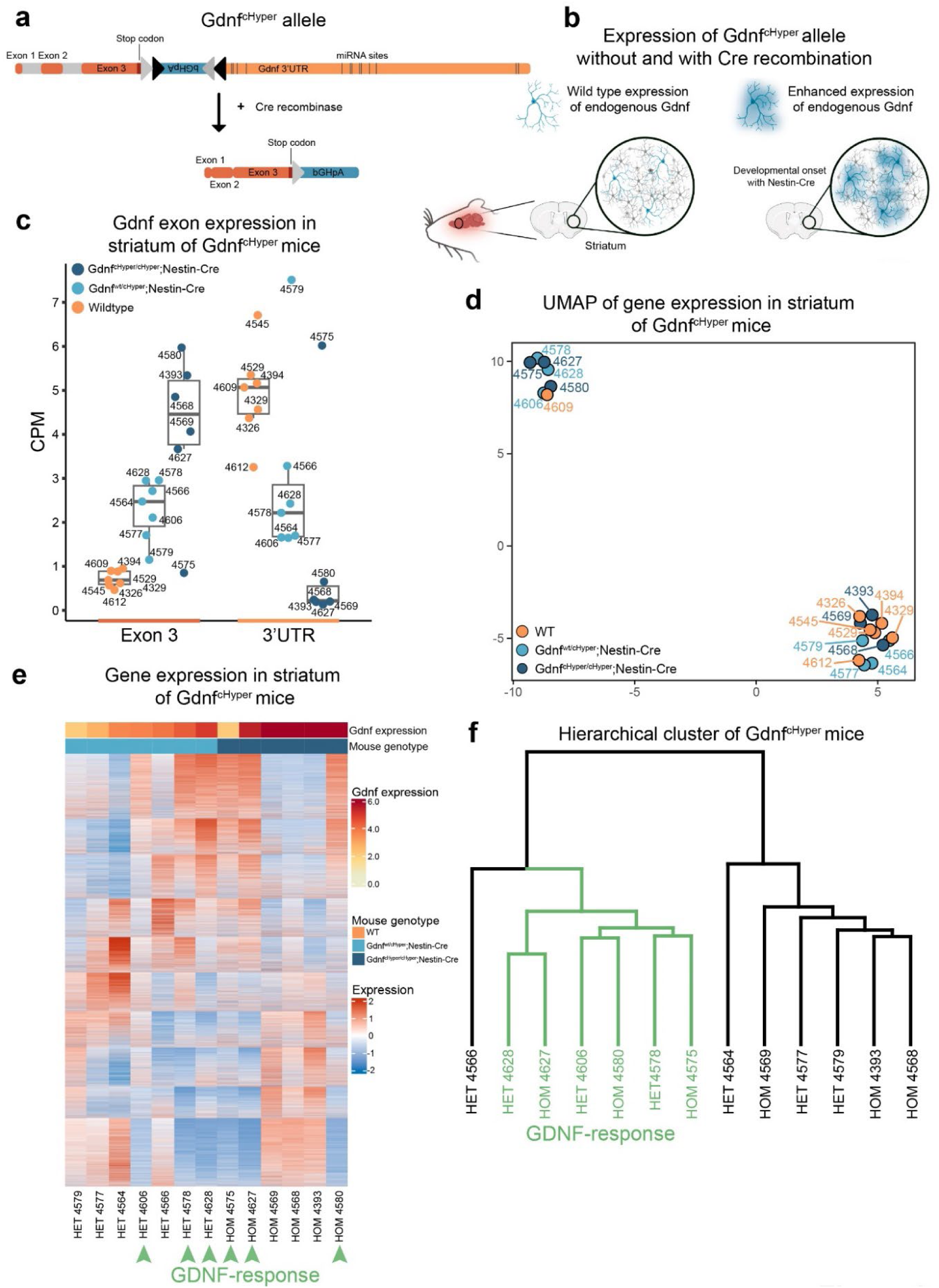
Gene expression patterns in Gdnf^cHyper^;Nestin-Cre mice with increased endogenous Gdnf. (**a**) Schematic of Gdnf^cHyper^ allele in Gdnf^cHyper^ mouse model and the resulting short mRNA after Cre recombination. (**b**) Schematic of enhanced Gdnf expression in cells naturally expressing Gdnf in STR in Gdnf^cHyper^ mice before and after recombination at mid-gestation around E11-E12 with Nestin-Cre. Created with BioRender.com. (**c)** Analysis of exon 3 and the natural 3’UTR sequence tags of Gdnf in heterozygous and homozygous Gdnf^cHyper^;Nestin-Cre animals compared with WT. (**d**) UMAP based on gene expression in STR of Gdnf^cHyper^;Nestin-Cre. Three out of six heterozygous Gdnf^wt/cHyper^;Nestin-Cre mice and three out of seven homozygous Gdnf^cHyper/cHyper^;Nestin-Cre mice cluster separately (upper left corner). (**e**) Gene expression pattern in STR in Gdnf^cHyper^;Nestin-Cre mice. The same six mice that clustered differently in D display a different gene expression pattern; these are labeled “GDNF-response”. (**f**) Hierarchical clustering of Gdnf^cHyper^;Nestin-Cre mice based on gene expression show that the GDNF-response mice form a separate cluster.

Next, we investigated gene expression patterns among *Gdnf^cHyper^*;Nestin-Cre animals and hypothesized that heterozygous *Gdnf^wt/cHyper^*;Nestin-Cre animals and homozygous *Gdnf^cHyper/cHyper^*;Nestin-cre animals would cluster separately. Interestingly, we found that, contrary to our hypothesis, in UMAP of gene expressions, three *Gdnf^wt/cHyper^*;Nestin-Cre mice and three *Gdnf^cHyper/cHyper^*;Nestin-cre, as well as one WT mouse, clustered separately from the rest (Fig. 2d, individual animals denoted by identifying numbers). Further investigation via a heatmap of the gene expression patterns revealed that these six *Gdnf^cHyper^*;Nestin-Cre animals that clustered separately displayed a different gene expression pattern (Fig. 2e). In hierarchical clustering, we find them to form a separate cluster (Fig. 2f). We labeled these animals “GDNF-response” mice. In a differential gene expression analysis on this group, we found 2414 significantly upregulated genes and 2760 significantly downregulated genes compared with WT (FDR < 0.05; list of genes in Extended Data Table 4). Since mature GDNF protein encoding exon 3 is increased in allele-dose dependent manner in all animals as expected (Fig. 2c), these data suggest that GDNF-response gene expression pattern in STR is a physiological state that does not ubiquitously happen in all animals.

### Comparative striatal gene expression in SCZ patients and GDNF-response mice

To increase our understanding of GDNF in SCZ, we compared the striatal gene expression between SCZ patients and *Gdnf^cHyper^*;Nestin-Cre mice. Using the DEGs from GDNF-response mice, a comparative heatmap revealed the most similarities in gene expression patterns between the GDNF-response mice and the four GDNF-high SCZ patients (Fig. 1c-d) that stand out from the rest of the patients (Extended Data fig. 3a). Comparison of DE genes in GDNF-response mice and the four GDNF-high SCZ patients revealed 1534 downregulated genes (Fig. 3a, P = 4.1e^−294^) and 389 upregulated genes (Fig. 3b, P = 0.99; Extended Data Table 5). Thus, we found strong similarities between downregulated genes but not among upregulated genes. Based on these results, we label these four GDNF-high patients with SCZ as “GDNF-response” patients.

**Fig. 3.**
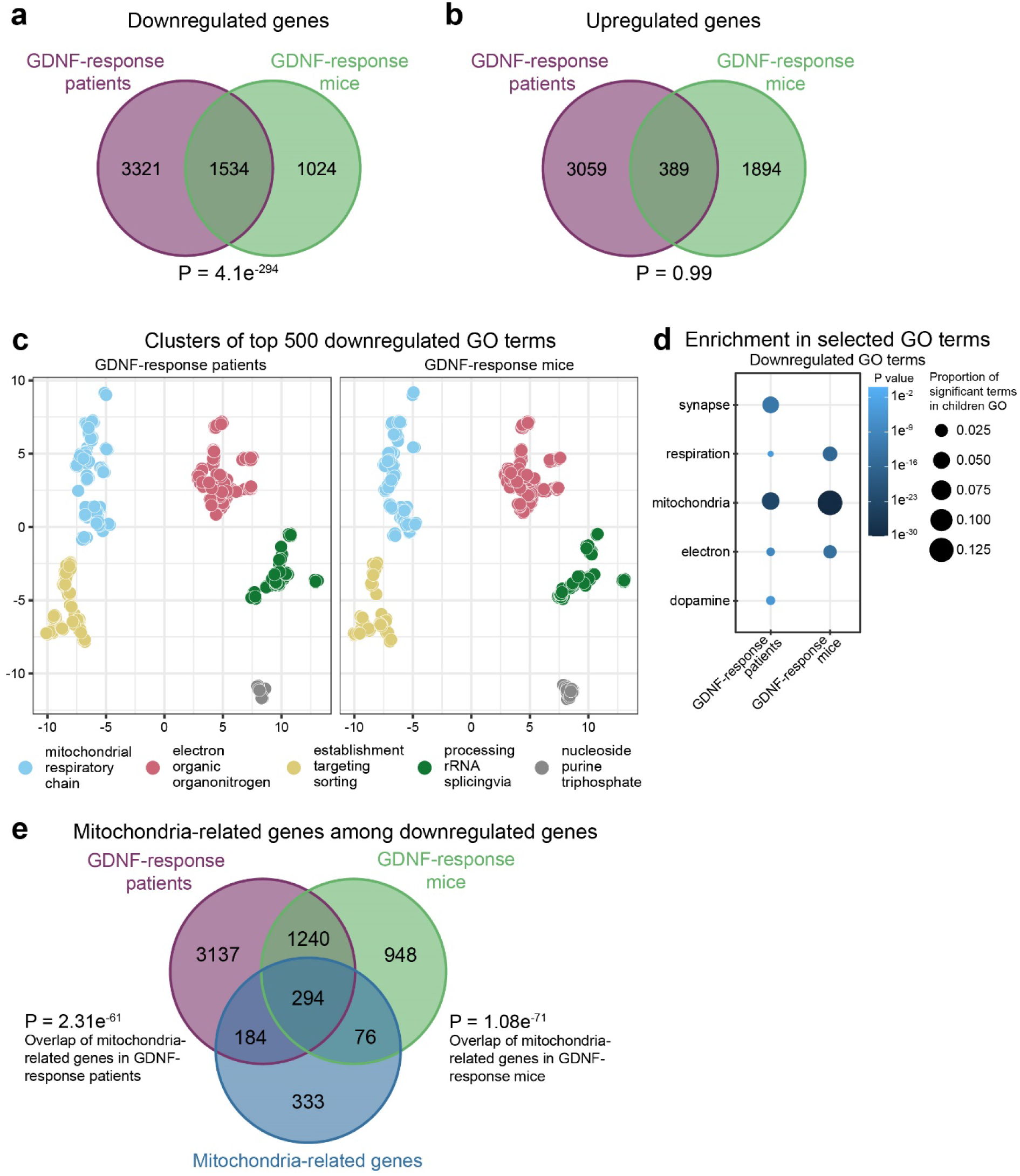
Comparable striatal downregulation in a subgroup of patients with SCZ and GDNF-response mice. (**a,b**) Overlapping downregulated (a) and upregulated (b) genes between GDNF-response patients and GDNF-response mice (FDR < 0.05). Significant overlap among (P = 4.1e^−294^; one-sided hypergeometric test) among downregulated genes, but not for upregulated genes. (**c**) UMAP clustering of the top 500 most significant GO terms from downregulated genes in GDNF-response patients and GDNF-response mice, respectively. (**d**) Enrichment of selected GO terms in downregulated GO terms in GDNF-response mice and GDNF-response patients. Only significant terms are shown. (**e**) Significant enrichment of mitochondria-related genes among downregulated genes in both GDNF-response patients (P = 2.31e^−61^; one-sided hypergeometric test) and GDNF-response mice (P = 1.08e^−71^; one-sided hypergeometric test).

Next, we investigated the functionality of the DEGs by performing gene ontology (GO) analysis, focusing on biological processes. We explored the enrichment of GO terms (P < 0.01) at level 2 for the two GDNF-response groups and found that in both patients and mice, *metabolic process*, *localization,* and *cellular process* are the largest GO groups originating from downregulated genes (Extended Data Fig. 3b; Extended Data Table 6 and 7). Next, we used semantic similarity in UMAP to cluster the top 500 downregulated terms (Fig. 3c). We named the clusters with three of the most prominent words appearing in the GO terms. The largest clusters from downregulated GO terms in both GDNF-response groups relate to *mitochondria*, *respiration*, and the *electron chain*, as well as *nucleosides*, *sorting*, and *processing RNA* (Fig. 3c). Thereafter, we focused our attention on specific GO terms among downregulated genes and used recurring words to sort the GO terms into relevant functional groups, relating to synapses, respiration, mitochondria, electrons, and DA (Fig. 3d). We observed that GO terms relating to mitochondria were most significant in both, thus, we inspected mitochondria-related genes closely (Fig. 3e) and found enrichment among downregulated genes in both GDNF-response patients (478 genes, P = 2.31e^−61^) and GDNF-response mice (370 genes, P = 1.08e^−71^). Further analysis using Kyoto Encyclopedia of Genes and Genomes (KEGG) revealed that one of the most affected pathways in both GDNF-response patients and GDNF-response mice was *oxidative phosphorylation*, which occurs in the inner mitochondrial membrane (Extended Data Fig. 4). We found downregulation in all five complexes (I-V) in both groups (Extended Data Fig. 4a-b).

### Striatal GDNF expression negatively correlates with expression of DA-related genes in PFC

In *Gdnf^cHyper^*;Nestin-Cre mice, PFC tissue DA levels are reduced, while cyclic voltammetry analysis of dopamine release and reuptake in PFC projecting dopaminergic ventral tegmental area (VTA) is decreased^29^ (Fig. 4a). Similar analysis in patients is not possible; hence, we turned to data on post-mortem gene expression. Unfortunately, two patients (SCZ 8 and 15) out of the four GDNF-response patients were missing gene expression data from PFC in the current data set^30^, making a gene expression comparison of PFC genes and their relation to striatal GDNF expression levels statistically unreliable. Therefore, we analyzed correlations between DA-related genes in PFC and the expression of striatal GDNF in all available patient samples. We found that about twice as many DA-related genes were negatively correlated (31 genes) with striatal GDNF as were positively correlated (16 genes; Fig. 4b-c; P < 0.1). Next, we visualized the correlation between all DA-related genes in PFC and striatal GDNF in the KEGG pathway *dopaminergic synapse* (Fig. 4d, Extended Data Fig. 5). Here, we observe that high striatal GDNF associates with downregulation of PFC DA transmission – excitatory post-synaptic D1 receptor level was negatively correlated, while the D2 receptor, which primarily inhibits DA signaling as an autoreceptor, was positively correlated with striatal GDNF level. VMAT, which transports DA from the cytoplasm to synaptic vesicles prior to release, was negatively correlated, while PFC DA-degrading enzymes catechol-*O*-methyltransferase (COMT) and monoamine oxidase (MAO) expression levels were positively correlated with striatal GDNF levels. Taken together, these findings indicate a link between high striatal GDNF and decreased DA signaling in the PFC.

**Fig. 4.**
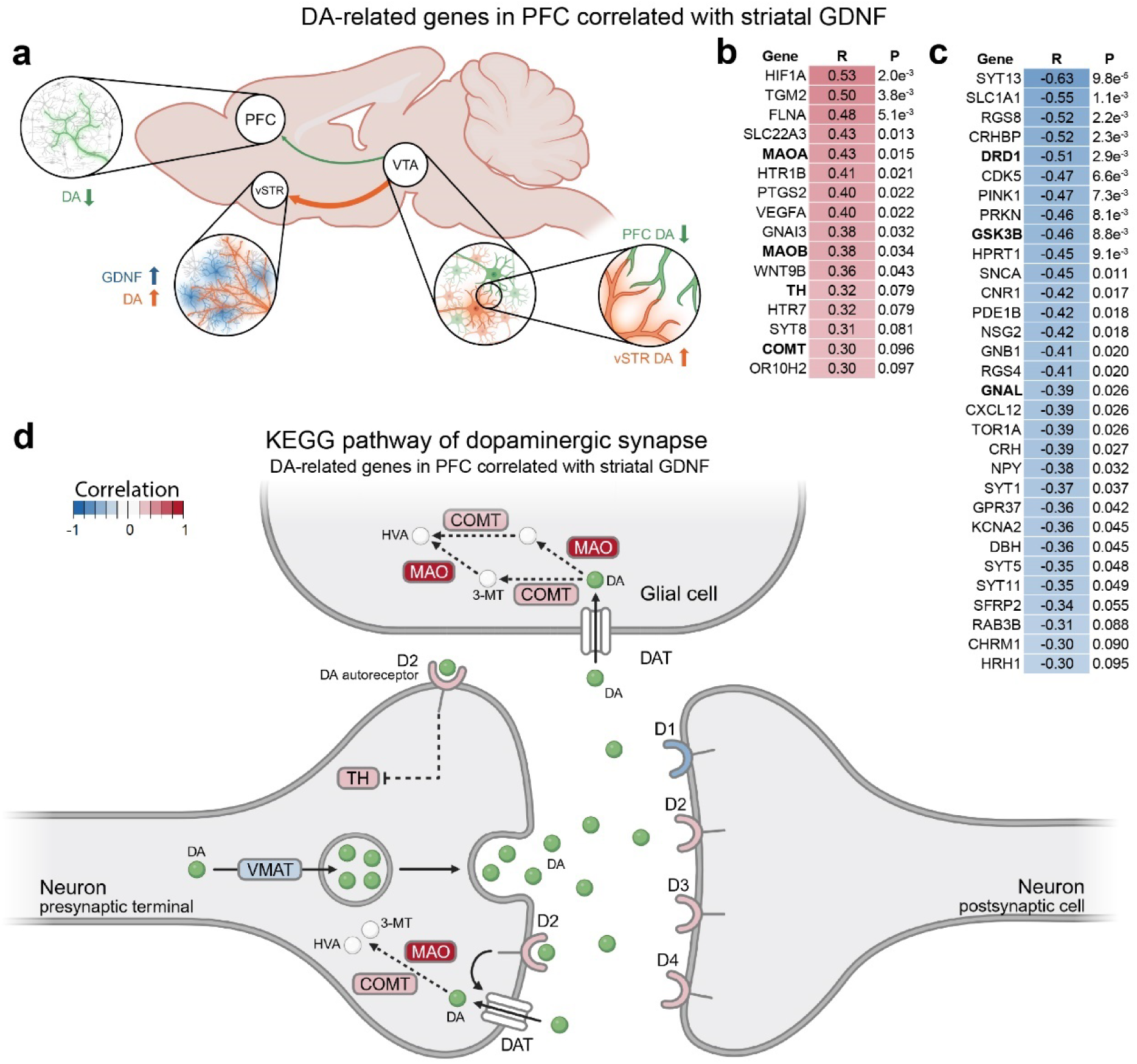
Correlation between striatal GDNF and DA in PFC. (**a**) DA neurons in VTA project to ventral STR (vSTR) and PFC in mice. Endogenous upregulation of striatal Gdnf leads to increased striatal DA but decreased DA signaling in PFC ^29^. Created with BioRender.com. (**b-c**) In post-mortem PFC of SCZ patients and healthy controls, there are 31 DA-related genes that correlate negatively (c) with striatal GDNF and 16 DA-related genes that correlate positively (b) (P < 0.1; Pearson’s correlation). (**d**) Simplified schematics of KEGG pathway of dopaminergic synapse with DA-related genes in PFC correlated with striatal GDNF in SCZ patients and healthy controls. Created with BioRender.com.

### GDNF in CSF of FEP patients correlates with CSF DA and clinical test scores

As a next step, we analyzed two independent cohorts of patients with FEP and HCs using CSF, serum, and clinical test scores – data from KaSP FEP patient cohort (Sweden) and LMU Hospital FEP cohort (Germany). Starting with patients with FEP in the KaSP, we’ve assessed the correlation of the levels of GDNF, DA, and other neurotransmitters and neuronal function regulators in the CSF. We found a correlation between GDNF and DA, but no correlation with other neurotransmitters or neuronal function regulators (Fig. 5a, P = 2.8e^−4^). The correlation between GDNF and DA in CSF was statistically significant in patients with FEP (Fig. 5b; P = 1.6e^−4^), but a trend was also seen in HCs (P = 0.058). Furthermore, correlation analyses of CSF GDNF and symptom ratings in patients revealed that CSF GDNF levels correlated with PANSS negative scores (Fig. 5c; P = 0.054), but not with PANSS positive scores (Extended Data Fig. 6a; P = 0.47). The PANSS general scores show a trend towards correlation with CSF GDNF (Extended Data Fig. 6b; P = 0.15), and taken together, this results in a positive correlation trend for total PANSS scores and CSF GDNF (Extended Data Fig. 6c; P = 0.091). In line with previous report^29^, there was a positive correlation between CSF GDNF and Clinical Global Impression (CGI) scores (P = 0.015; Extended Data Fig. 6d). In patients with FEP in the cohort from LMU Hospital we found similar significant correlations between CSF GDNF (Fig. 5d-f) and PANSS negative scores (Fig. 5d; P = 0.041) and PANSS general scores (Fig. 5e; P = 0.0014), and we observed a trend towards positive correlation in PANSS positive scores (Extended Data Fig. 6e). In LMU FEP cohort, the total PANSS scores correlated with GDNF levels in the CSF (Extended Data Fig. 6f). Similar to the KaSP patient cohort, we observed a correlation between CSF GDNF and CGI scores in the LMU Hospital patients with FEP (Fig. 5f; P = 0.026). Thus, elevated CSF GDNF correlates with high CSF DA levels and with poorer outcome in tests measuring negative symptoms and overall disease scores.

**Fig. 5.**
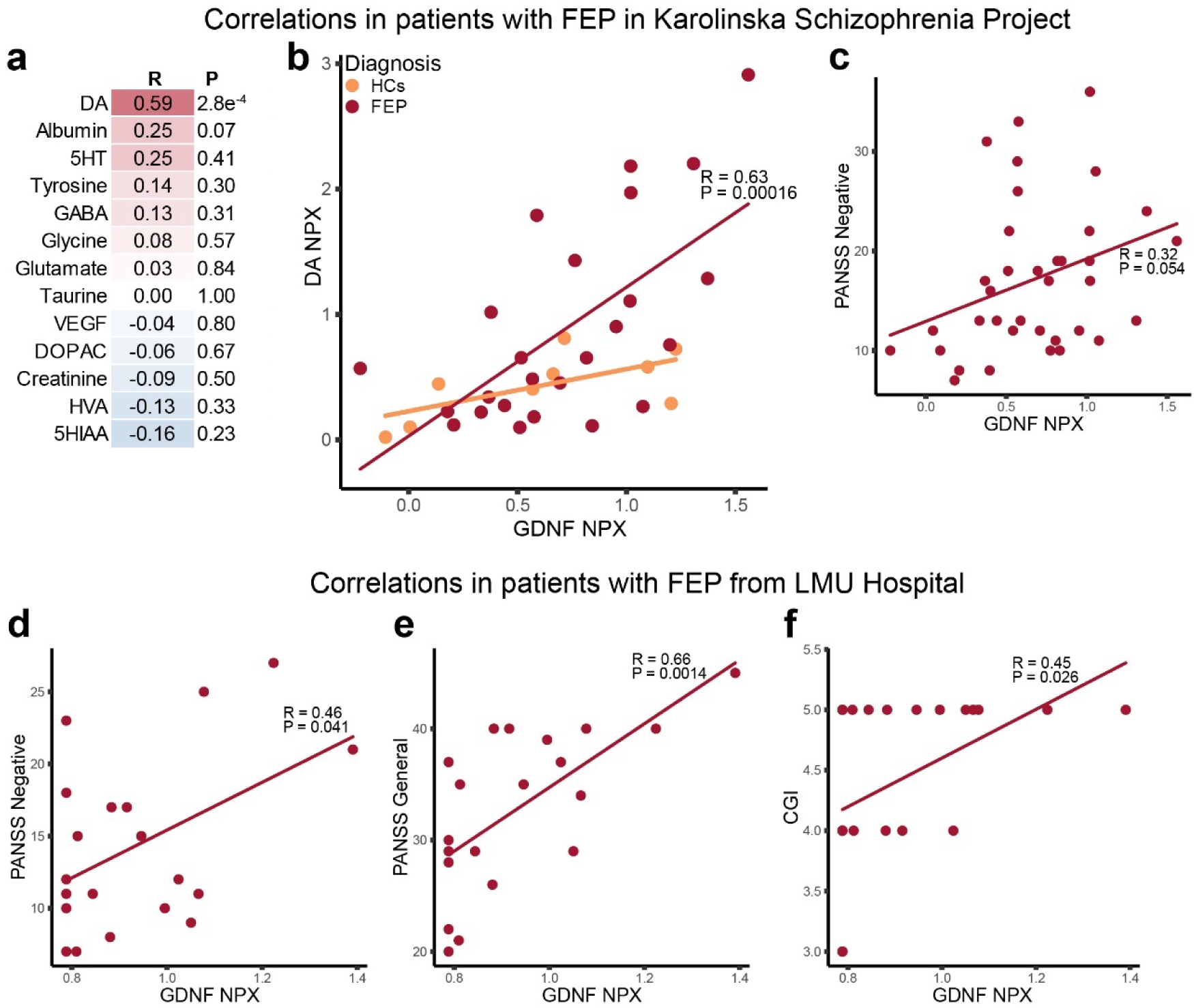
Correlations between CSF GDNF and clinical test scores in two independent cohorts of FEP patients: Karolinska Schizophrenia Project (KaSP; a-c) and LMU Hospital (d-f). (**a-b**) Correlation between neurotransmitter levels and GDNF in CSF in FEP patients from KaSP and healthy controls (Pearson’s correlation) (a). DA levels are positively correlated with GDNF levels in both patients with FEP and HCs, respectively (b). (**c**) PANSS negative scores positively correlate with GDNF in CSF in FEP patients in KaSP (P = 0.054). (**d-f**) Positive correlations between CSF GDNF in FEP patients from LMU Hospital and PANSS Negative scores (d; P = 0.041), PANSS General scores (e; P = 0.0014), and CGI scores (f; P = 0.026).

## DISCUSSION

GDNF is among the strongest dopaminergic neurotrophic factors known and has therefore gained wide attention as a potential therapeutic for treating Parkinson’s disease (PD), where striatal DA is depleted and enhancement of the remaining striatal DA fibers is needed^24,28,33,34^. On the other hand, DA-enhancing drugs, including psychostimulants and DA agonists used to alleviate PD symptoms, increase the risk of psychosis at higher doses^35,36^. Notably, some of these drugs, including methamphetamine, also increase nigrostriatal GDNF expression in several mammalian species^37–39^. In the current study, we found elevated GDNF expression in STR in seven out of eighteen patients with SCZ and identified a gene expression pattern that we termed “GDNF-response” in four of the patients with SCZ with elevated GDNF in the STR. Interestingly, we found a similar GDNF-response pattern in about half of *Gdnf^cHyper^*; Nestin-Cre mice with increased endogenous striatal GDNF. Notably, the two GDNF-response groups – patients with SCZ and *Gdnf^cHyper^*; Nestin-Cre mice – share more than 1500 significantly downregulated genes (P = 4.1e^−294^). We observed enrichment in mitochondria-related genes among these (P = 2.31e^−61^ and P = 1.08e^−71^, respectively), including downregulation in the pathways of oxidative phosphorylation, previously reported to associate with SCZ^40–49^. Why a uniform GDNF-response is not observed in all GDNF-high mice and patients remains unknown. It may reflect interindividual stochastically emerging differences in feedback loops, such as in RET tyrosine kinase inhibitor SPROUTY 1 and SPROUTY2, PTPN11/SHP2 phosphatase and SOCS activity levels, or in similar inhibitory pathways further downstream the signaling cascade via PI3K/AKT/mTOR and MAPK systems^50–57^.

In SCZ, increased DA signaling in the STR is likely causing the positive symptoms of psychosis, while decreased DA signaling in PFC is believed to contribute to cognitive dysfunction and negative symptoms such as apathy. *Gdnf^cHyper^*;Nestin-Cre mice with about 2-fold increase of GDNF in the central nervous system (CNS) reflect the human DA signaling patterns in SCZ – increased DA in the STR and decreased DA in the PFC – and display SCZ-like symptoms^29^. Our current results from patient samples align with these findings. The main source of brain DA (∼90%) is the STR, which is the only site in the brain where DA neurons are in direct and in *en masse* contact with GDNF-expressing neurons^22,23,58,59^. Thus, it is likely that the observed correlation between GDNF and DA levels in CSF in patients with FEP reflects increased striatal GDNF expression, which led to increased striatal DA production that is a well-established function of striatal GDNF in PD models and unlesioned animals^26,27,29,60–63^. Unfortunately, methods enabling direct analysis of tissue DA levels in the PFC of live FEP patients are not available. However, analysis of FEP patients revealed that higher CSF GDNF correlated with stronger general and negative symptoms in SCZ. In line with the important relationship of higher GDNF with the individual disease burden, analysis of post-mortem STR and PFC samples revealed negative correlations between striatal GDNF and DA function and transmission-related gene expression in the PFC.

Importantly, DA neurons respond to GDNF only via the RET receptor^64–66^, and GDNF is not expressed in the neurons in the PFC^22,23,58^. GDNF stimulates striatal DA production^26,27,29,60–63^. Thus, the most likely scenario for disease mechanism in GDNF-response patients and GDNF-response mice is that striatal GDNF overproduction directly stimulates RET-bearing, nigrostriatal and mesolimbic DA fibers, which originate from the substantia nigra (SN) and VTA (Fig. 6). VTA also contains PFC-projecting mesocortical DA neurons^67,68^. It is likely that mesolimbic VTA neurons that overproduce DA deliver paracrine signals to neighboring PFC-projecting mesocortical DA neurons, instructing them to reduce DA production. This, in turn, leads to downregulation of PFC DA metabolism as observed in GDNF-response patients and mice with developmental and adult onset striatal increase in GDNF production^29^, summarized in (Fig. 6).

**Fig. 6.**
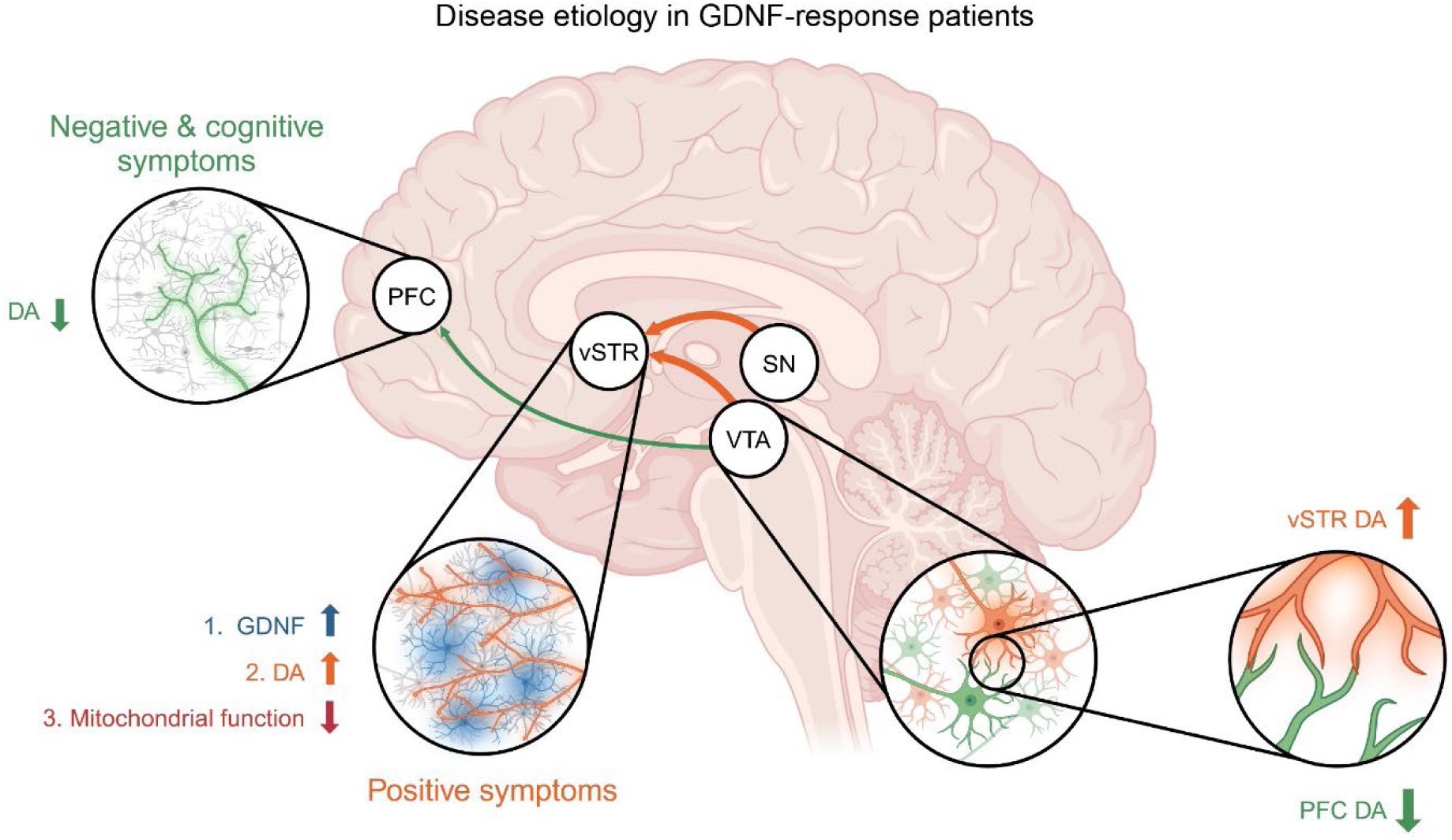
Hypothesized disease etiology in GDNF-response patients. In GDNF-response patients, GDNF is upregulated in ventral STR (vSTR), which leads to increased DA signaling but decreased mitochondrial function. Dopaminergic neurons originating in the VTA project to both the vSTR and the PFC, suggesting that the VTA can become a site for communication between STR-projecting and PFC-projecting dopaminergic neurons. Likely, the STR-projecting neurons instruct the PFC-projecting neurons to decrease their DA production, leading to decreased DA signaling in the PFC, and negative and cognitive symptoms arise in the patients. Created with BioRender.com.

Collectively, our data suggest that enhanced GDNF expression may be prominently involved in the disease etiology in about 20% of patients with SCZ. Thus, the development of a drug that inhibits elevated GDNF signaling may allow for addressing a disease-driving mechanism, and its application in GDNF-high FEP subgroup of patients may block disease progression into SCZ. Pharmacological inhibition of GDNF signaling may also reduce side effects associated with DA receptor blocking antipsychotic drugs since expression of GDNF and its main receptor RET is restricted to just a few neuronal types in the CNS^23,58,64,65,69^. For example, the tuberoinfundibular DA pathway (TIDA system) projects from the hypothalamus into median eminence and negatively regulates prolactin secretion from the pituitary gland. Related side effects are often referred to as among the most life-quality reducing side-effects of DA-blocking drugs. Those include increased breast growth and lactation in both genders, loss of libido, and increased body weight. These side effects may be avoided by putative GDNF system antagonists, since GDNF is not expressed in the projection targets of those neurons^22,23,58^. Furthermore, in comparison to nigral and VTA DA neurons, where almost 100% of DA neurons express RET, at least in mice^70^, RET expression in tuberoinfundibular DA neurons is low or absent^58,71^. Targeting GDNF signaling is likely to dampen DA metabolism specifically in the STR, as earlier genetic work with striatal AAV-Cre delivery into GDNF conditional Knock-Out mice suggests^72^. As previously demonstrated in mice^29^ and reported in patients in this study, excess in striatal GDNF associates with reduction in PFC DA metabolism. Pharmacological inhibition of GDNF could therefore constitute pharmacological route to treat low PFC DA-related negative features.

Silencing of the striatal presynaptic DA system via GDNF inhibition may not only be applicable as precision medicine to treat GDNF-high patients, but may also treat disease as a stand-alone or auxiliary drug in all patients with elevated striatal DA, which make up about 50% of all SCZ patients^9,11,12,20^.

Taken together, we believe our work provides evidence of a novel disease-driving mechanism, candidate biomarker, and possible therapeutic target for a subgroup of SCZ patients, paving the way for future applied research across multiple areas.

## MATERIALS AND METHODS

### Data accession

Raw microarray data from human post-mortem brain samples were obtained from Lanz et al. ^30^ and accessed via GEO: GSE53987. We have previously published RNAseq data from Gdnf^cHyper^ mice^29^ and the data is accessible at GEO: GSE162974.

### Pre-processing and differential gene expression analysis

Microarray data of human post-mortem patient samples was imported to R and pre-processed using oligo (RRID:SCR_015729) followed by annotation with affycoretools^74^. Differential gene expression analysis was performed with edgeR (RRID:SCR_012802) and with additional linear modelling fitting with function eBayes in limma^75^. Genes without annotated Entrez IDs were excluded. For the full SCZ patient group, FDR = 0.1 was used as cut-off limit, whereas for the analysis separating the GDNF-response SCZ patients, FDR = 0.05 was used.

Fastq files from RNAseq of *Gdnf^cHyper^* mice were aligned to mouse reference genome mm10 with STAR (RRID:SCR_004463). Mapped reads in annotated exons were counted by featureCounts (RRID:SCR_012919), and the result was imported to R. Mapped exons and the 3’UTR of *Gdnf* was counted separately, and the counts from exons (natural 3’UTR excluded) were used for differential gene expression analysis. Genes with at least one count per million (CPM) in at least three samples were used in differential gene expression analysis using edgeR. Genes were normalized with TMM normalization, trended with dispersion estimation. The significance cut-off was set to FDR = 0.1 for *Gdnf^wt/cHyper^*;Nestin-Cre and *Gdnf^cHyper/cHyper^*;Nestin-Cre animals and FDR = 0.05 for GDNF-response mice. Plotting of GDNF exon count was done with ggplot2^73^.

### GDNF expression in post-mortem SCZ patients

Volcano plot of SCZ patient gene expression and boxplot of GDNF expression was plotted with ggplot2^73^. The GDNF expression was transformed via linear regression (1.31m-7.475) based on Fig. 2 in Zhao et al.^31^ to more closely resemble RNAseq data, and transformed with –0.278543 for centering so mean of Ctrl = 0.

### Defining GDNF-response subgroup in SCZ patients and Gdnf^cHyper^;Nestin-Cre mice

UMAPs of gene expression in patients and *Gdnf^cHyper^*;Nestin-Cre mice were supervised by disease or genotype, and performed with uwot^74^. UMAPs were plotted with ggplot2^73^. Heatmaps of patient gene expression were created with ComplexHeatmap (RRID:SCR_017270). Genes were som-clustered and expression values standardized to mean equals 0 and standard deviation equals 1. Hierarchical clustering of *Gdnf^cHyper^*;Nestin-Cre mice was performed with *hclust* in R, using method ward.D2. Dendrograms were created with the package factoextra (RRID:SCR_016692).

### Comparing gene expressions between data sets

A list of homologues with entrez ids and gene symbols for human and mouse genes was created using NCBI’s Ensembl annotations, and this list was utilized when comparing between data sets of the different species. Genes without homologues were excluded. Significance for overlapping DE genes between data sets were tested with one-sided hypergeometric tests. Venn diagrams of the overlaps were built using VennDiagram (RRID:SCR_002414).

### Functional analysis of GO terms

GO analysis was performed with *goana* in limma^75^, and GO terms with P < 0.01 was determined significant. The data base GO.db (RRID:SCR_002811) was used for relating GO terms to each other and assigning levels. In GO term enrichment analysis, only significant GO terms were used (P < 0.01). Representing the selected GO term groups relating to synapses, respiration, mitochondria, electrons, and dopamine, GO terms were grouped by their terms containing “synap”, “respi”, “mitoc”, “electron”, and “dopa” respectively. ggplot2^73^ was used for visualization of GO term enrichment and one-sided hypergeometric testing was used to determine the significance, with P < 0.01 set as significant. For creating UMAPs showing GO term similarity, top 250 (upregulated) or top 500 (downregulated) GO terms were used for semantic similarity calculations with GOSemSim^76,77^ (Wang’s method), mapped with uwot^74^, and clustered with dynamicTreeCut^78^. Word count was used for naming the clusters and the clustering of the GO terms was visualized with ggplot2^73^. Mitochondria-related and dopamine-related genes were determined by translating mitochondria-related and dopamine-related GO terms, respectively, back to genes using the database org.Hs.egGO2ALLEGS. Chord diagrams were created using the package circlize^79^ in R. GOSemSim^76,77^ was used for testing semantic similarity between GO term groups, using Wang’s method, and visualized with ggplot2^73^.

### KEGG pathways

KEGG diagrams were made with Pathview (RRID:SCR_002732). The oxidative phosphorylation diagrams were created with differentially expressed genes in pathway hsa00190, with logFC guiding the coloring. Bacteria– and archaea-specific genes were excluded from the diagrams and headings introduced for clarification. The dopaminergic synapse diagram was rendered using DA-related genes in PFC correlated with striatal GDNF in pathway hsa04728, correlation coefficient as color guide.

### CSF and serum patient samples from LMU Hospital in Munich

A total of 47 (37 males, 10 females) individuals with SSDs treated at the Department for Psychiatry and Psychotherapy of the LMU Hospital, Munich, Germany, were enrolled between 16.07.2021 and 13.03.2023. Inclusion criteria comprised age between 18 and 60 years and a diagnosis of SSD, according to DSM-V, assessed with the Mini International Neuropsychiatric Interview (M.I.N.I.), German version 7.0.2^80^. This includes the following diagnoses: schizophrenia, schizoaffective disorder, and brief psychotic disorder. All were inpatients, included at various stages of their disorder.

Exclusion criteria were defined as follows: a primary psychiatric disorder other than SSD, current electroconvulsive therapy or non-invasive brain stimulation treatment, coercive treatment, acute suicidality, any central nervous system (CNS) disorder other than SSD, history of traumatic brain injury (TBI), severe somatic (e.g., inflammatory, rheumatic) diseases, acute infections, current pregnancy (excluded in clinical routine) or lactation, regular current drug abuse (in the past month), inability to provide informed consent, current participation in clinical trials. Prior to inclusion in the study, all participants provided written informed consent. The study protocol was approved a priori by the local ethics committee of the LMU Munich (reference numbers 21-1139 and 21-0183). The study was conducted according to the Declaration of Helsinki.

CSF was collected via lumbar puncture at either the L4-L5 or L5-S1 interspace. In order to avoid carry-over effects from serum proteins, samples with erythrocyte counts above 500/µL were excluded. The CSF samples were processed within an hour by first centrifuging at 4°C at 400 x g for 10 min, followed by discarding the pellet of cells and debris. The supernatant was placed in – 80°C for storage prior to proteomic analysis.

Serum samples were collected at the same time point as CSF samples. Sample tubes were inverted 5 times directly after collection, and thereafter the blood were allowed to clot undisturbed at room temperature for 20-30 min, followed by centrifugation at 4°C at 2000 x g for 10 min. The supernatant was placed in –80°C for storage prior to proteomic analysis. The CSF samples were analyzed in the laboratory of our hospital by a cell-based assay from EUROIMMUN (Lübeck, Germany) for autoimmune encephalitis (antibodies tested: NMDAR, α-amino-3-hydroxy-5-methyl-4-isoxazolepropionic acid receptor-1 (AMPAR-1), α-amino-3-hydroxy-5-methyl-4-isoxazolepropionic acid receptor-2 (AMPAR-2), CASPR2, LGI1 and γ-amino-butyric acid receptor B1/B2 [n = 25]). No antibodies were detected in CSF, and only one subject manifested NMDAR antibodies in serum.

### CSF patient samples from the Karolinska Schizophrenia Project in Stockholm

CSF from 39 FEP patients (23 males and 16 females, ages 18-44 years) was collected via the Karolinska Schizophrenia Project (KaSP), with an ethical permit from Stockholm Regional Ethics Committee, Dnr 2010/879-31/1. FEP patients were recruited between 2011 and 2019, at four psychiatric clinics located in Stockholm, Sweden. All patients were examined by at least two board-certified psychiatrists, and clinical assessment was done with Structured Clinical Interview for DSM-IV Axis I Disorders, PANSS, the GAF scale, CGI, NAB Mazes, LNS, and a Fluency test. Patients with other neurological diseases, severe somatic illness, or drug addiction were excluded.

Healthy controls were recruited via advertisement and underwent physical examination, screening of blood and urine, and MRI examination. Psychiatric and psychotic illness, as well as use of illegal drugs, were exclusion criteria. CSF from 21 healthy controls was used, 10 males and 11 females, ages 20-36 years.

CSF was collected between 7:45 AM and 10:00 AM, following a night of rest. Individuals were placed in the right decubitus position, and a disposable atraumatic needle (22G Sprotte, Geisingen, Germany) was inserted at the L4-5 level. From each individual, 18 ml CSF was collected. The samples were centrifuged at 1438g for 10 minutes at 4 °C (Sigma 5810R, Eppendorf, Hamburg, Germany at 3500 r.p.m.) and the supernatants were stored at –80°C. High-performance liquid chromatography (HPLC) for measuring neurotransmitters and metabolites was performed as previously described^81^. DA was measured in 9 healthy controls and 24 FEP patients.

### Measurement of GDNF in patient CSF samples using Olink Proximity Extension Assay

GDNF in human CSF patient samples were measured with either the panel “Inflammation” (KaSP patient samples) or the panel “Target 96 Neurology” (LMU patient samples) from Olink Bioscience (Uppsala, Stockholm). The assays use multiple proximity extension assay (PEA) technology, where protein markers are measured in microtiter plates with DNA-labeled antibody probe pairs. Target protein binding to the antibody will make the PEA probes draw nearer and initiate DNA polymerization. A new DNA sequence is formed, representing the target protein. The DNA sequence is quantified with real-time PCR. The data was processed using the R package OlinkAnalyze (RRID: SCR_022438). The results were normalized for inter– and intra-run variation using internal– and interplate controls, and processed data is presented in log2-scale as Normalized Protein eXpression (NPX).

### Correlation analysis between GDNF and neurotransmitters, metabolites, and clinical test scores in patients with FEP

Pearson’s correlation coefficient was used to determine correlations between GDNF in CSF and other neurotransmitters and neuronal function regulators, or test scores in clinical tests.

Correlation graphs were created with ggplot2^73^.

### Protein analysis of patient serum samples using LC-MS/MS

Protein samples were digested using a standard in-solution tryptic digestion protocol. Peptides were desalted using C18-based solid-phase extraction before LC-MS/MS analysis. Peptide mixtures are analyzed on a Thermo Scientific Orbitrap Astral mass spectrometer coupled to an Evosep One nanoLC system. Peptides are separated on a 15 cm C18 column using a pre-programmed Evosep 60 samples per day (SPD) method, with a total gradient length of ∼21 minutes. The Astral is operated in data-independent acquisition (DIA) mode, using optimized scan settings for high-speed acquisition and deep proteome coverage. DIA data were analyzed using DIA-NN with a human UniProt spectral library. Protein quantification was based on precursor-level quantification, and data normalization was performed using median normalization.

To assess individual classification performance of each protein, a univariate analysis was applied by calculating the area under the curve (AUC) for the receiver operating characteristics (ROC) curve for each protein using pROC^82^. The ROC curve represents the trade-off between true positive and false positive rates across varying decision thresholds, with higher AUC values indicating superior discriminative ability. Proteins with high AUC values, the top 5%, were used in principal component analysis (PCA) and UMAP (performed with the package umap^83^) to visualize separation between patients. Plots were created with ggplot2^73^ and ggpubr^84^. Additionally, to assess the classification accuracy based on the selected biomarkers, we implemented a multi-class ROC analysis using a support vector machine (SVM) model using e1071^85^.

## Data Availability

The code used for the analysis is available upon request. CSF from LMU is available upon reasonable request and will require MTA/DTA agreements.

## Acknowledgments

We would like to thank BEA, the Bioinformatics and Expression Analysis core facility, which is supported by the board of research at the Karolinska Institute. During this study, JOA, IR and AD have been supported by Center of Innovative Medicine (CIMED), Hjärnfonden, Team Rynkeby-God Morgon Skolloppet, Åhlén-stiftelsen, Swedish Research Council (grants no. 2019-01578 and 2022-01093). In addition, JOA and AM have been funded by the Academy of Finland (grants no. 297727, JOA and 350678, JOA), Sigrid Juselius Foundation, ERA-NET NEURON grant nr 352077, JOA and AM, Helsinki Institute of Life Science Research Fellow, JAES Foundation grant no. 240034, JOA, and by HORIZON-RIA grant nr 101188432 DTRIP4H, to JOA. DRG has been funded by University of Helsinki Doctoral Programme Brain and Mind. Furthermore, this research was supported by the Federal Ministry of Education and Research (Bundesministerium für Bildung und Forschung [BMBF]) with the EraNet project GDNF UpReg (01EW2206) to PF, AS, and VY. The study was endorsed by the BMBF within the initial phase of the German Center for Mental Health (DZPG) (grant: 01EE2303A, 01EE2303F to PF, AS). The study was funded by the Supplement to BMBF funding for the German Centre for Mental Health (DZPG) by the Bavarian State Ministry for Science and the Arts with the Grant for the research project ‘Improving Infrastructures for DZPG and NAKO Cohorts” to PF. The study was funded by the EU HORIZON-INFRA-2024-TECH-01-04 project DTRIP4H 101188432 to PF, AS and FR. VY was supported by the Residency/PhD track of the International Max Planck Research School for Translational Psychiatry (IMPRS-TP). VY was supported by the Faculty of Medicine at LMU Munich (FöFoLe Reg.-Nr. 1226/2024). JM was supported by the Faculty of Medicine at LMU Munich (FöFoLe Reg.-Nr. 1167). FJR received funding from the Pesl-Alzheimer-Stiftung (2024-2025), from the Lisa Oehler-Stiftung (2022–2024) and from the Verum-Stiftung (2024-2025). C.M.S (KaSP cohort) was supported by grants from Erling Persson Family Foundation, Swedish Research Council (2023-02827), Swedish Society for Medical Research (CG-23-0298-B), and Region Stockholm (FoUI-986509).

## Author contributions

Conceived and designed the study: JOA, IR, DG, AM, AD.

Performed the experiments: IR, DG, AM, VI, SO, SK, MV.

Analyzed the data: IR, DG, JOA, MM, AM, FO, VY, JM, SK, MV.

Performed bioinformatics analysis: IR, AD, PC, MM, FBB.

Supervised bioinformatics analysis: AD.

Collected KaSP patient cohort: GE, CS, SC, SE.

Provided KaSP cohort related data: GE, FO.

Collected LMU FEP cohort and provided related data: CDP-Workingroup, VY, JM, AS, PF, EW, FR.

Funding: JOA, FR, PF, AS, EW.

Wrote the paper: IR, JOA.

All co-authors read and contributed to the writing of the paper.

## Data and materials availability

The code used for the analysis will be available at GitHub prior to publishing. CSF from LMU is available upon reasonable request and will require MTA/DTA agreements.

