## Extended Data for "Excess GDNF associates with a subtype of schizophrenia with enhanced dopamine and increased prefrontal dysfunction"

### Extended Data Figures

#### Extended Data Figure 1

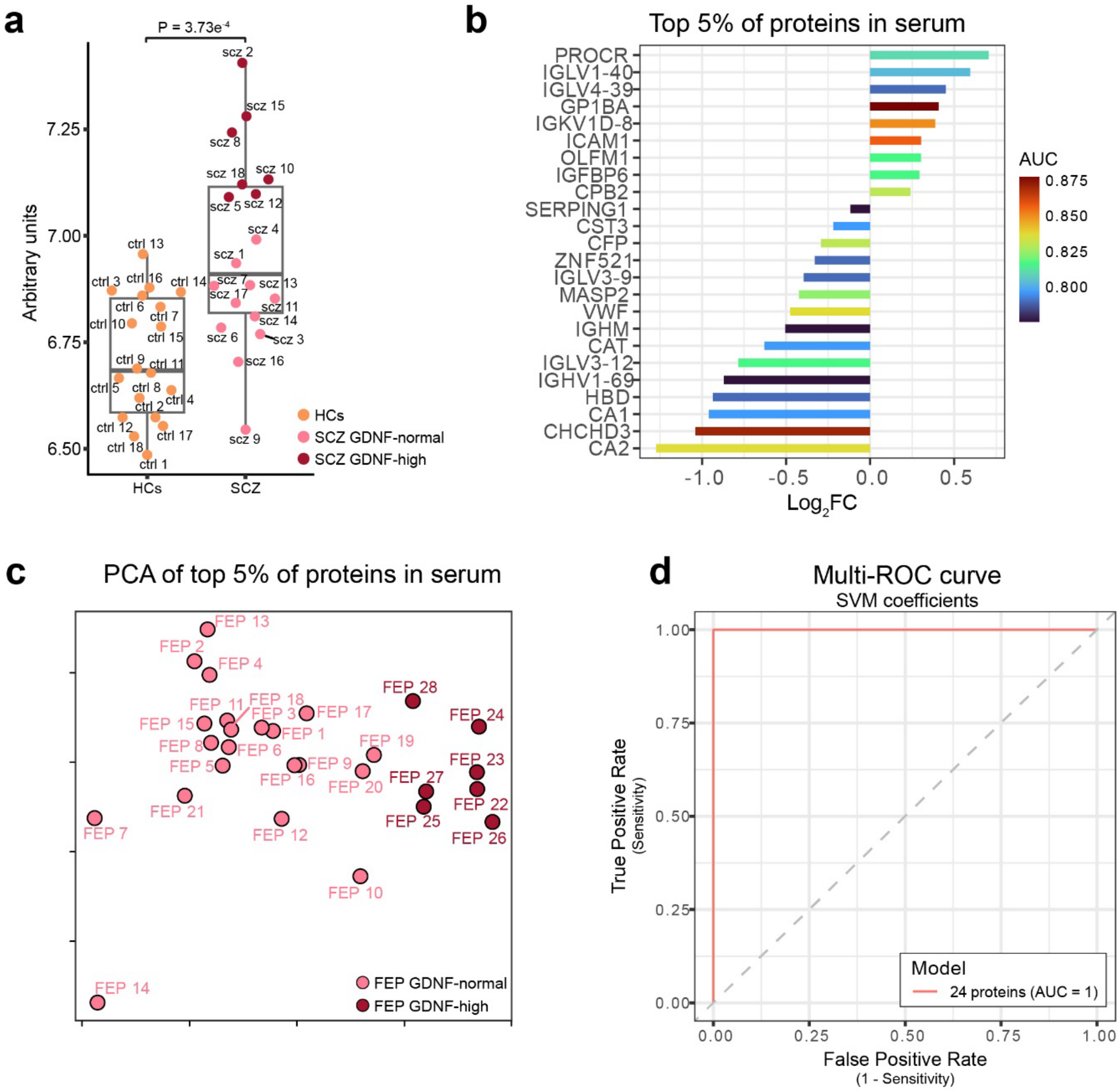


**Extended Data Fig. 1. GDNF expression in post-mortem STR in patients with SCZ and HCs and protein analysis in serum from patients with FEP.** (**A**) Untransformed GDNF expression levels in patients with SCZ (N=18) and HCs (N=18) from microarray data. Based on raw data of postmortem STR patient samples[30]. (**b-d**) Protein analysis in serum samples from patients with FEP. (**b**) The 24 proteins, and their $Log_2$ fold changes, corresponding to the top 5% proteins with the highest AUC in individual univariate analysis. (**c**) PCA based on the top 5% proteins in (B), with patients with FEP are colored based on their GDNF level in CSF. Patients with high GDNF tend to separate from patients with normal GDNF levels. (**d**) Multi-ROC curve with SVM coefficients using the top 5% proteins as a model gives a classification accuracy of AUC = 1.

**Extended Data Figure 2**

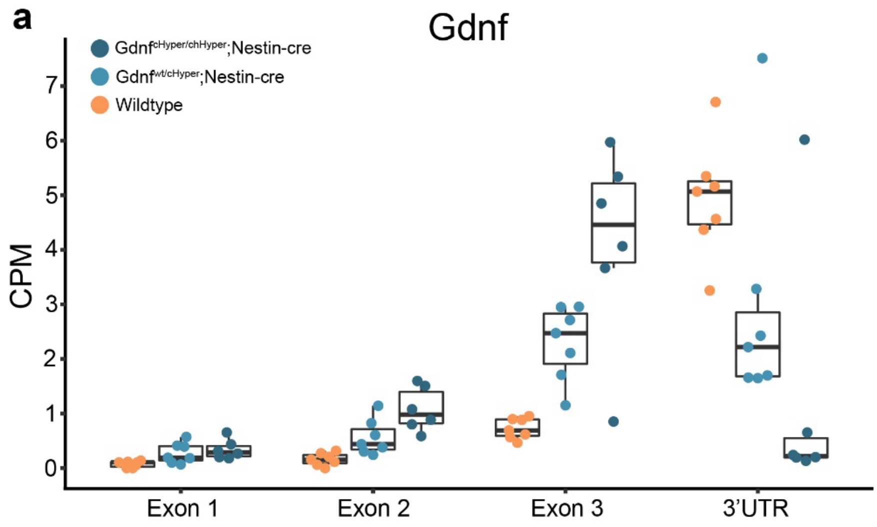


**Extended Data Fig. 2. Exon expression of Gdnf in Gdnf$^{cHyper}$ mice.** Expression of exons 1-3 and the natural 3'UTR sequence tags of Gdnf in heterozygous and homozygous Gdnf$^{cHyper}$;Nestin-Cre animals compared with WT[29].

### Extended Data Figure 3

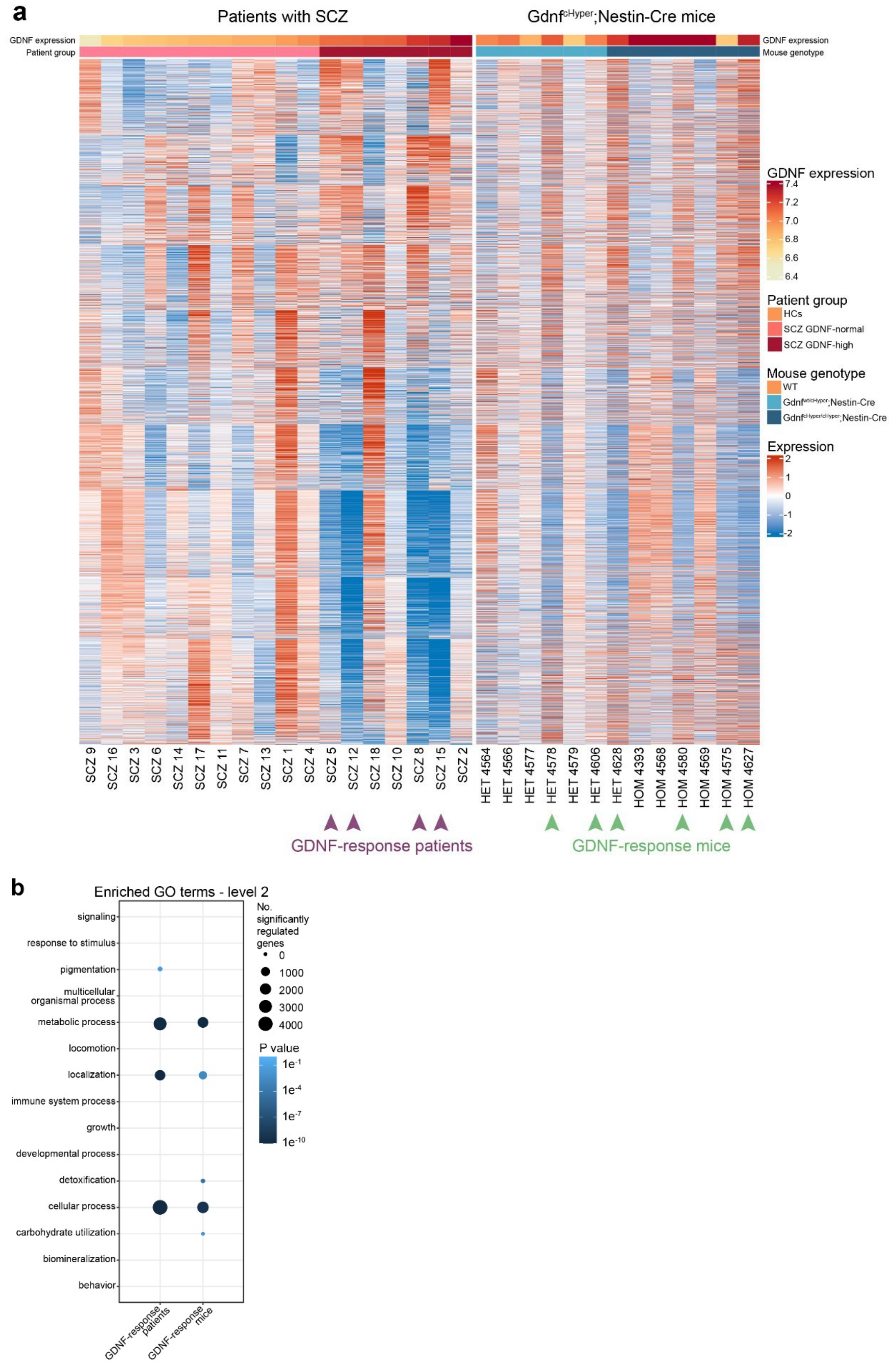

**Extended Data Fig. 3. Gene expression pattern similarities and GO term level 2 enrichment.** (**a**) Heatmap of genes DE in GDNF-response mice, shown for individual patients and mice. GDNF-response patients and GDNF-response mice are marked with arrows. (**b**) Enrichment among level 2 GO terms from downregulated genes in GDNF-response patients and GDNF-response mice. Only significant ($P < 0.01$) GO terms are shown.

**Extended Data Figure 4**

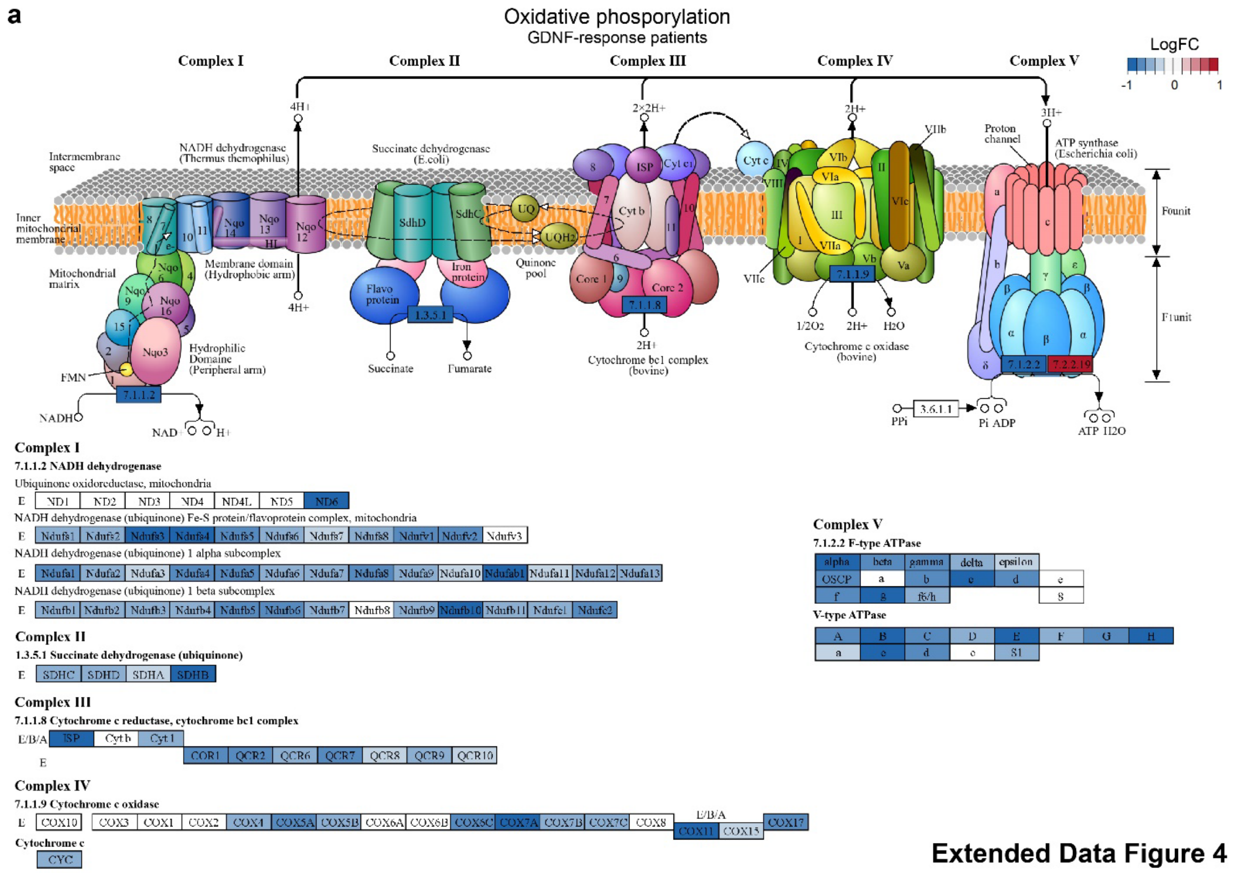

b

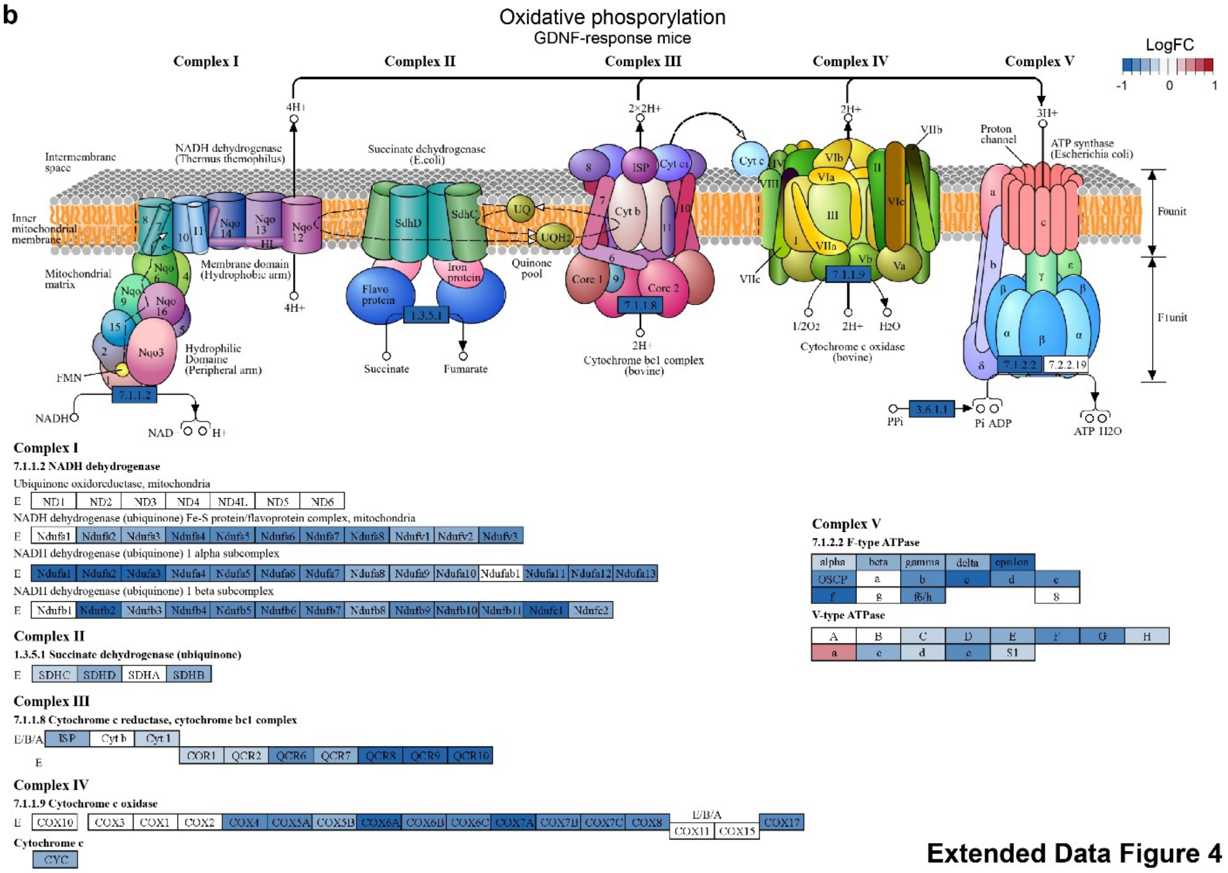


Extended Data Figure 4

**Extended Data Fig. 4. Regulation of oxidative phosphorylation in GDNF-response groups.**

(**a-b**) Regulation of differentially expressed genes (FDR < 0.05) in GDNF-response patients (a) and GDNF-response mice (b) in the KEGG pathway Oxidative Phosphorylation. Only mammalian genes are shown. Genes are colored based on logFC; blue = downregulation and red = upregulation.

**Extended Data Figure 5**

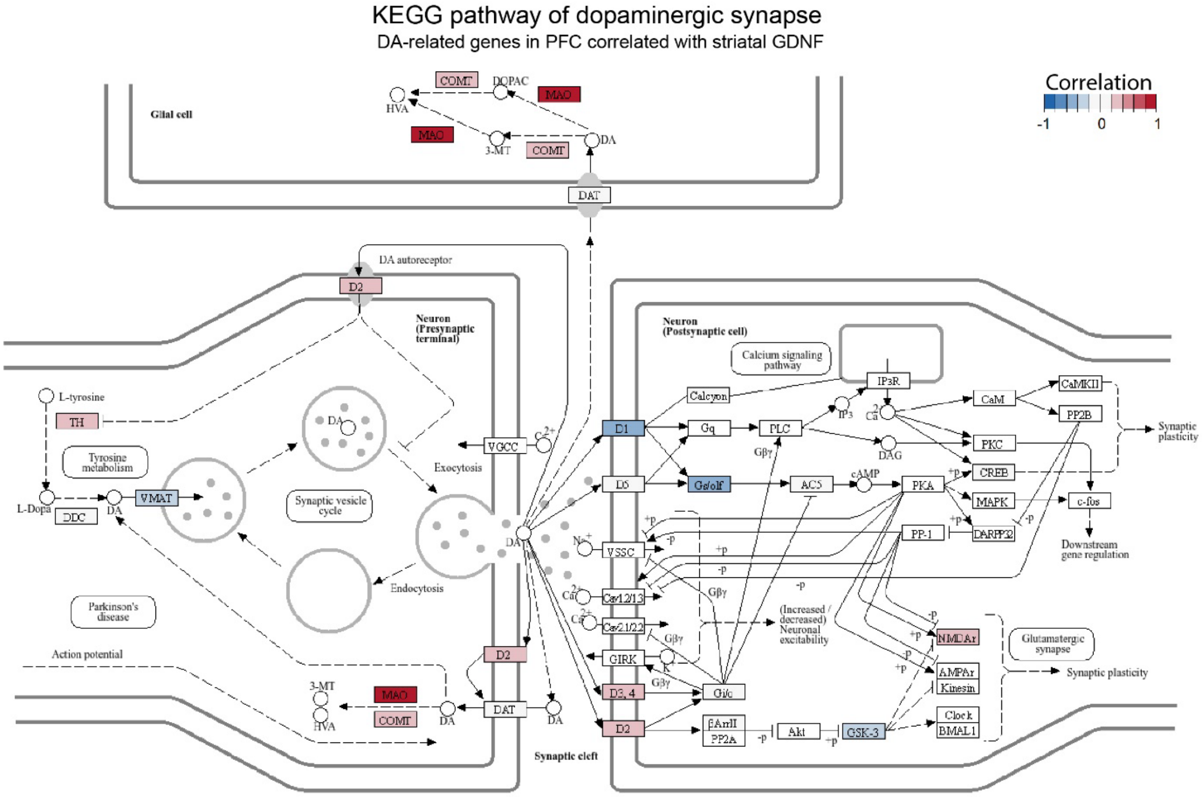


**Extended Data Fig. 5. Genes DA synapse in PFC correlated with striatal GDNF.** Full KEGG pathway of dopaminergic synapse with DA-related genes in PFC correlated with striatal GDNF in patients with SCZ.

**Extended Data Figure 6**

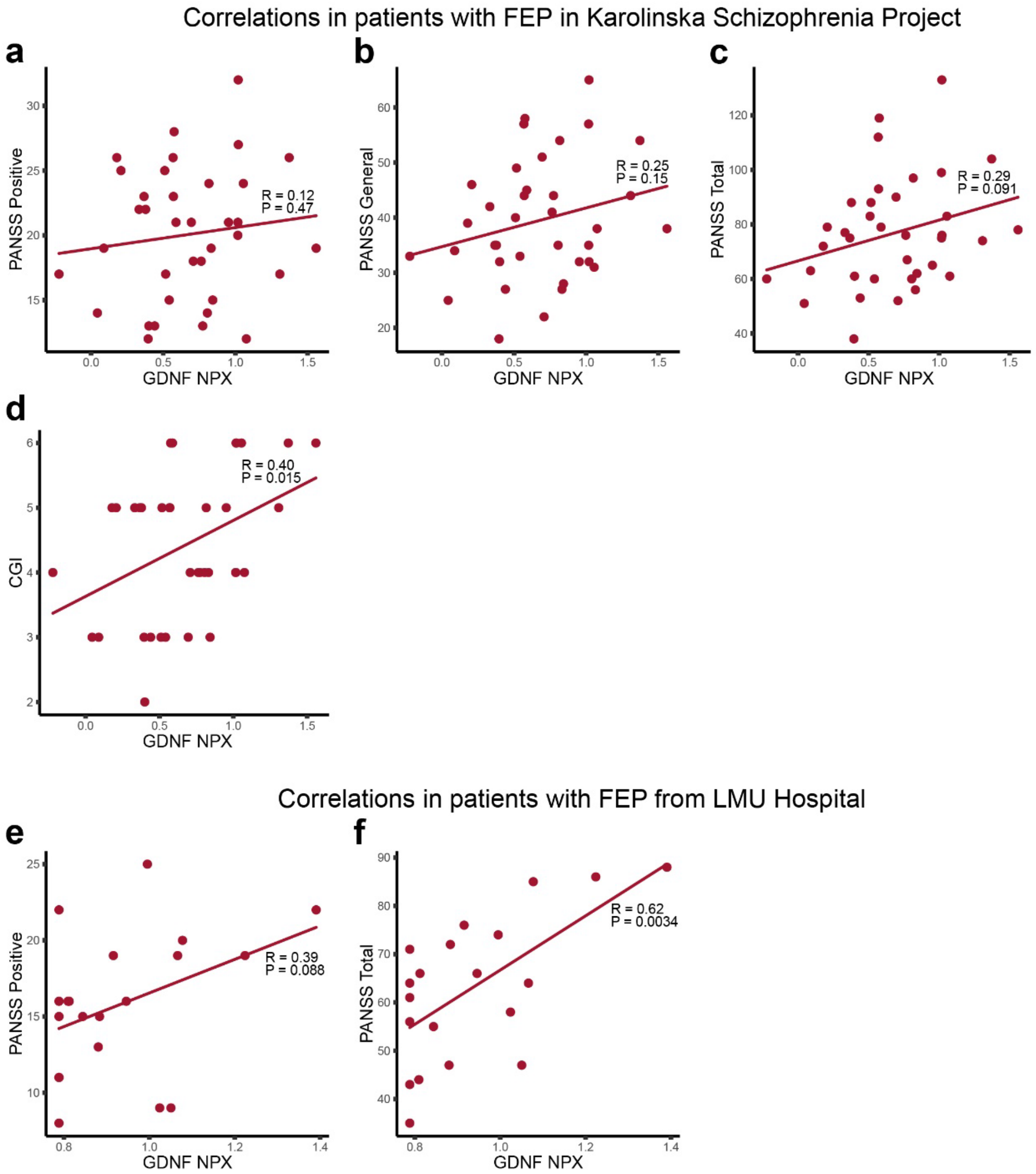


**Extended Data Fig. 6. Correlation of CSF GDNF and clinical test scores in two independent FEP patient cohorts.** (**a-c**) CSF GDNF in patients with FEP in KaSP does not correlate with PANSS positive scores (a; P = 0.47) but shows a trend towards significant correlation with PANSS general (b; P = 0.15) and PANSS total scores (c; P = 0.091). (**d**) Correlation between Clinical Global Impression (CGI) scores and CSF GDNF in patients with FEP in KaSP (P = 0.015). (**e-f**) CSF GDNF in patients with FEP from LMU

Hospital show a trend towards positive correlation with PANSS positive scores (e; P = 0.088), and a significant correlation with PANSS total scores (f; P = 0.0034).

#### Extended Data Tables

Attached as separate files.

**Extended Data Table 1.** Differentially expressed genes in patients with SCZ.

**Extended Data Table 2.** Differentially expressed genes in GDNF-response patients.

**Extended Data Table 3.** Differentially expressed genes in $Gdnf^{cHyper}$ mouse models.

**Extended Data Table 4.** Differentially expressed genes in GDNF-response mice.

**Extended Data Table 5.** Differentially expressed genes in common between GDNF-response patients and GDNF-response mice.

**Extended Data Table 6.** Differentially expressed GO terms in GDNF-response patients.

**Extended Data Table 7.** Differentially expressed GO terms in GDNF-response mice.
